# Detection and genotyping of enteric foodborne viruses with high (NoV, HAV, HEV) and low (RV, AdV) health impact in ready-to-eat leafy vegetables and berries from Córdoba, Argentina

**DOI:** 10.1101/2025.02.26.25322975

**Authors:** Guadalupe Di Cola, Verónica E. Prez, Anabella C. Fantilli, Camila Frydman, Marina Mozgovoj, Liliana Luque, Leonardo Ferreyra, Silvia V. Nates, María Belén Pisano, Viviana E. Ré

**Affiliations:** Instituto de Virología “Dr. J. M. Vanella” - Facultad de Ciencias Médicas - Universidad Nacional de Córdoba, Argentina; Consejo Nacional de Investigaciones Científicas y Técnicas (CONICET), Argentina; Instituto Nacional de Tecnología Agropecuaria (INTA), Instituto Tecnología de Alimentos, Hurlingham, Argentina; Instituto de Ciencia y Tecnología de Sistemas Alimentarios Sustentables, UEDD INTA CONICET, Hurlingham, Argentina; Departamento Laboratorio Central, Ministerio de Salud de la Provincia de Córdoba, Argentina

**Keywords:** enteric viruses, foodborne, vegetables, berries, Argentina

## Abstract

The global consumption of ready-to-eat (RTE) leafy green vegetables and berries has risen as consumers perceive them as safe and nutritious options. However, these foods have also been identified as sources of enteric viruses that infect the human gastrointestinal system, which are then excreted and can spread through the fecal-oral route. In Argentina, there is limited evidence on the detection of enteric viruses in food, and no legislation currently requires their detection in frozen or fresh produce intended for domestic consumption. This study aimed to identify and characterize five foodborne viruses with high and low health impact (norovirus, hepatitis A virus, hepatitis E virus, rotavirus, and adenovirus) in berries and RTE leafy vegetables marketed in central Argentina. A total of 242 samples (145 berries and 97 leafy greens) were collected from retail sources in Córdoba and processed according to ISO 15216-2:2019 standard. Viral detection was performed by qPCR or RT-qPCR, followed by viral characterization through specific RT-PCR assays or Sanger sequencing. Results indicate that 4.1% of berry samples, primarily frozen strawberries, tested positive for norovirus or rotavirus, while 10.3% of leafy green samples were positive for norovirus, rotavirus and adenovirus. Genotyping indicated the same strains to those involved in local gastroenteritis outbreaks. The absence of hepatitis A and E viruses suggests limited transmission of these pathogens through these foods. This study highlights the need to strengthen hygiene and monitoring protocols for RTE products, as viral contamination poses potential health risks to consumers, underscoring a critical area for improving food safety.

## 1. Introduction

Urbanization and changes in food consumer habits have increased the number of people buying and eating ready-to-eat (RTE) foods. Globalization has triggered growing consumer demand for a wider variety of foods, resulting in an increasingly complex and longer global food chain. Access to adequate and safe food is key to sustaining life and promoting good health. Unsafe food, contaminated with harmful microorganisms or chemical substances, has been implicated in over 200 diseases, spanning from gastrointestinal issues to severe diseases (WHO, 2022).

The consumption of fruits and vegetables is widely recognized as part of a healthy diet. Soft fruits, such as berries with their delicate nature, are typically handpicked due to their sensitivity to handling and potential damage. After harvesting, these fruits often enter the market with minimal processing or are exported frozen without additional measures to eliminate pathogens (X. Gao et al., 2019). In the case of RTE green leafy vegetables, they undergo post-harvest processing, including decontamination with a sodium hypochlorite solution, to eliminate harmful microorganisms before being cut and packaged, making them available for consumption (Castro-Ibáñez et al., 2017). The implementation of Good Agricultural Practices, Good Manufacturing Practices and Good Hygiene Practices is imperative in minimizing the risk of microbial contamination and guaranteeing food safety throughout the entire production process (FAO/WHO, 2024). These systems help to identify potential hazards, establish control measures, and monitor critical points to ensure that fresh and frozen produce meet stringent safety standards before reaching consumers (Santos et al., 2023). However, these systems, as well as microbiological quality standards, focus mainly on bacterial control.

In recent years, there has been a growing concern about the role of viruses as causative agents of foodborne illnesses. Unlike bacteria, viruses do not replicate within food; however, they can contaminate foodstuffs and transmit infection (Seymour & Appleton, 2001). Viruses that infect the gastrointestinal tract are typically shed in feces. Despite shedding high viral loads, only a few viral particles are required to cause infection and subsequent illness (CAC, 2012). Food contamination can occur directly from infected individuals during handling, processing or through the environment, especially from irrigation water. While viruses primarily spread from person to person, epidemiological evidence suggests that they can be transmitted through minimally processed foods, including seafood, fruits, vegetables, and salads. The occurrence of foodborne outbreaks and illnesses associated with frozen and fresh products has raised concern regarding the safety of these consumable commodities (Nasheri et al., 2019). The Joint FAO/WHO Expert Meeting on Microbiological Risk Assessment of viruses in food (JEMRA) highlighted that the virus-commodity combinations ranked of highest priority were human norovirus (NoV) and hepatitis A virus (HAV) in fresh and frozen produce, prepared and RTE foods, and shellfish (FAO/WHO, 2023), as they are the primary vehicles of foodborne outbreaks (Di Cola et al., 2021; Tavoschi et al., 2015). In addition, other viruses are relevant to food safety, as they can contaminate food and water, and be transmitted to humans by these routes. This is the case of hepatitis E virus (HEV), which has also been recently highlighted by the JEMRA (FAO/WHO, 2024), rotavirus (RV), astrovirus, sapovirus and adenovirus (AdV) (Seymour & Appleton, 2001). Structurally, these viruses are non-enveloped, and they generally exhibit greater resistance to common control measures compared to bacteria, needing comprehensive strategies to ensure food safety (CAC, 2012).

Argentina is an agro-exporting country, with significant agricultural production. It annually produces over 58 thousand tons of soft fruits. The main provinces producing strawberries and blueberries are Tucumán (in the northern region), Santa Fe, Buenos Aires and Entre Ríos (Pampean region), with a smaller fraction produced in Patagonia (south region). A portion of these berries is earmarked for domestic consumption as fresh produce, while between 50% and 90% is exported frozen (RSA CONICET, 2016). Regarding green leafy vegetables, they have a larger production area across the country for domestic consumption.

Current local regulations concerning the identification of pathogenic microorganisms in plant-based foods primarily focus on detecting pathogenic bacteria, neglecting the identification of viruses (CAA, 2023), which constitutes a risk for consumers. The lack of regulation could be explained by: 1) scarce evidence (infrequently recognized as etiological agents of foodborne); 2) underestimation of the risk due to the health system’s lack of recognition of outbreaks caused by foodborne viruses; and 3) the historical idea that the presence of bacterial indicators (especially fecal coliforms) in food suggests potential viral risk, overlooking the fact that the absence of bacteria does not correlate with absence of viruses (Torok et al., 2019). In Argentina, there is limited research on the detection of enteric viruses in leafy vegetables and berries. A few previous studies have reported the presence of NoV and RV in post-harvested vegetables (Coluccini, 2020; Prez et al., 2018, 2021), as well as NoV in raspberries (Oteiza et al., 2022).

With this background, we aimed to provide data on virus contamination in food, a field little explored in our region by identifying and genetically characterizing foodborne enteric viruses with high (NoV, HAV and HEV) and low (RV and AdV) health impact in samples of RTE leafy green vegetables, as well as in fresh and frozen berries.

### 2. Methodology

### 2.1 Sample collection

A total of 242 samples, each consisting of 25 g of berries and leafy green vegetables obtained from retail stores in Córdoba City, the central area of Argentina, were analyzed. Fruit samples (n=145) came mainly from Tucumán, and to a lesser extent from the Pampean region. They were composed of strawberries (*Fragaria × ananassa*, n=113) and blueberries (*Vaccinium corymbosum,* n=42), gathered between October 2021 and January 2023. This included 102 samples of RTE frozen fruits (strawberries n=91, blueberries n=11) and 43 samples of fresh fruits (strawberries n=22, blueberries n=21), which require washing before consumption. For leafy green vegetables, a total of 97 samples were collected between April 2022 and July 2023. These samples, originating from regional farms in Córdoba province, comprised 72 RTE specimens of fresh rocket (*Eruca sativa*) and 25 of chicory (*Cichorium intybus*) commercialized in markets of the city. Fresh samples were refrigerated until analysis (within 48 hours), whereas frozen samples were stored at -20°C until processing.

### 2.2 Sample processing and nucleic acid extraction

Viral extraction from fruit and vegetable samples was conducted through elution, following the protocol described in the ISO 15216-2:2019 standard, using the bacteriophage PP7 of *Pseudomonas aeruginosa* as process control virus and to evaluate the efficiency (Poma et al., 2013; Rajal et al., 2007). The PP7 stock used had 1.05x10^8^ genomic copies per microliter (gc/µL). The procedure initiated with 25 g of berries or vegetables, coarsely chopped into approximately 2.5 cm x 2.5 cm x 2.5 cm pieces, transferred to a bag with a filter. Subsequently, 40 mL of tris-glycine-beef extract (TGBE) buffer pH 9.5 and PP7 were added, supplemented with 1140 U of *Aspergillus aculeatus* pectinase for berries. The mixture underwent incubation at room temperature with constant agitation at 60 rpm for 20 minutes, with pH monitoring and adjustment to 9.5 at 10-minute intervals for fruits, to allow viruses to be released from the surface of foods. The supernatant from the filtered compartment was clarified by centrifugation at 10,000 x *g* for 30 min at 4°C. The resulting supernatant was mixed and incubated for 60 minutes with a polyethylene glycol (PEG)/NaCl solution. After centrifugation (10,000 x *g* for 30 min at 4°C), the pellet was compacted (10,000 x *g* for 5 min at 4°C), and the supernatant was discarded. The pellet was resuspended in PBS, constituting the concentrate for nucleic acid extraction in the case of vegetables. For berries, an additional clarification step involving chloroform/butanol treatment and centrifugation (10,000 x *g* for 15 min at 4°C) was required before RNA extraction (ISO 15216-2:2019).

Nucleic acid extraction was carried out from 200 µL of the concentrated samples using the commercial kit SureFast® PREP DNA/RNA Virus (CONGEN), following the manufacturer’s instructions. The resulting 50 µL of eluate were stored at -80°C until DNA/RNA amplification.

### 2.3 Nucleic acid amplification

Enteric virus screening was carried out using real-time PCR (qPCR) or real-time RT-PCR (RT-qPCR). Berries were tested for NoV genogroup I (GI), NoV genogroup II (GII), RV, HAV and HEV. Additionally, in leafy green vegetable samples, AdV was evaluated. Primers and probes utilized for detecting NoV GI, NoV GII, HAV, HEV, RV, AdV and PP7 are detailed in **Table1** . The amplification process was conducted on a StepOne™ Real-Time PCR system from Applied Biosystems, employing TaqMan® Universal PCR Master Mix and TaqMan Fast Virus 1-Step Master Mix (Applied Biosystems), for DNA and RNA viruses, respectively. Samples that tested positive by RT-qPCR or qPCR underwent retesting via nested or heminested PCR for typing.

Nested or heminested-PCRs were carried out using GoTaq® DNA Polymerase (Promega, USA). For RNA viruses, the extracted nucleic acid was first reverse-transcribed into cDNA using *random hexamer primers* and ImProm-II Reverse Transcriptase (Promega, USA). The PCR products were separated by electrophoresis on a 2% agarose gel stained with SYBR™ Safe (Invitrogen) and visualized using a UVP transilluminator.

### 2.4 Quantification of PP7 and extraction efficiency

Extraction efficiency for each sample was expressed as the recovery rate, determined by the ratio of the number of PP7 copies recovered after RNA extraction to the total number of PP7 copies spiked into the sample. Recovered PP7 was quantified using a standard curve constructed from serial tenfold dilutions (ranging from 10^7^ to 10^3^ gc/μL) of a plasmid available in our laboratory, containing an amplified PP7 gene fragment. Extraction efficiency was expressed as the percentage of virus recovery. In instances where the recovery efficiency was less than 1%, 1/10 dilutions were conducted. Samples with recoveries of less than 1% were tested again from the beginning (ISO 15216-2:2019; Terio et al., 2017).

### 2.5 Viral characterization

The characterization of RV involved the amplification of VP4 and VP7 genes using cDNAs, followed by multiplex heminested-PCRs using genotype-specific primers for G genotypes (VP7) and P genotypes (VP4) (**Table 1** ), as described by Gentsch et al., (1992) and Gouvea et al., (1990). The resulting PCR products were separated using 10% polyacrylamide gel electrophoresis (Laemmli, 1970) and visualized by silver staining (Herring et al., 1982).

**Table 1.**
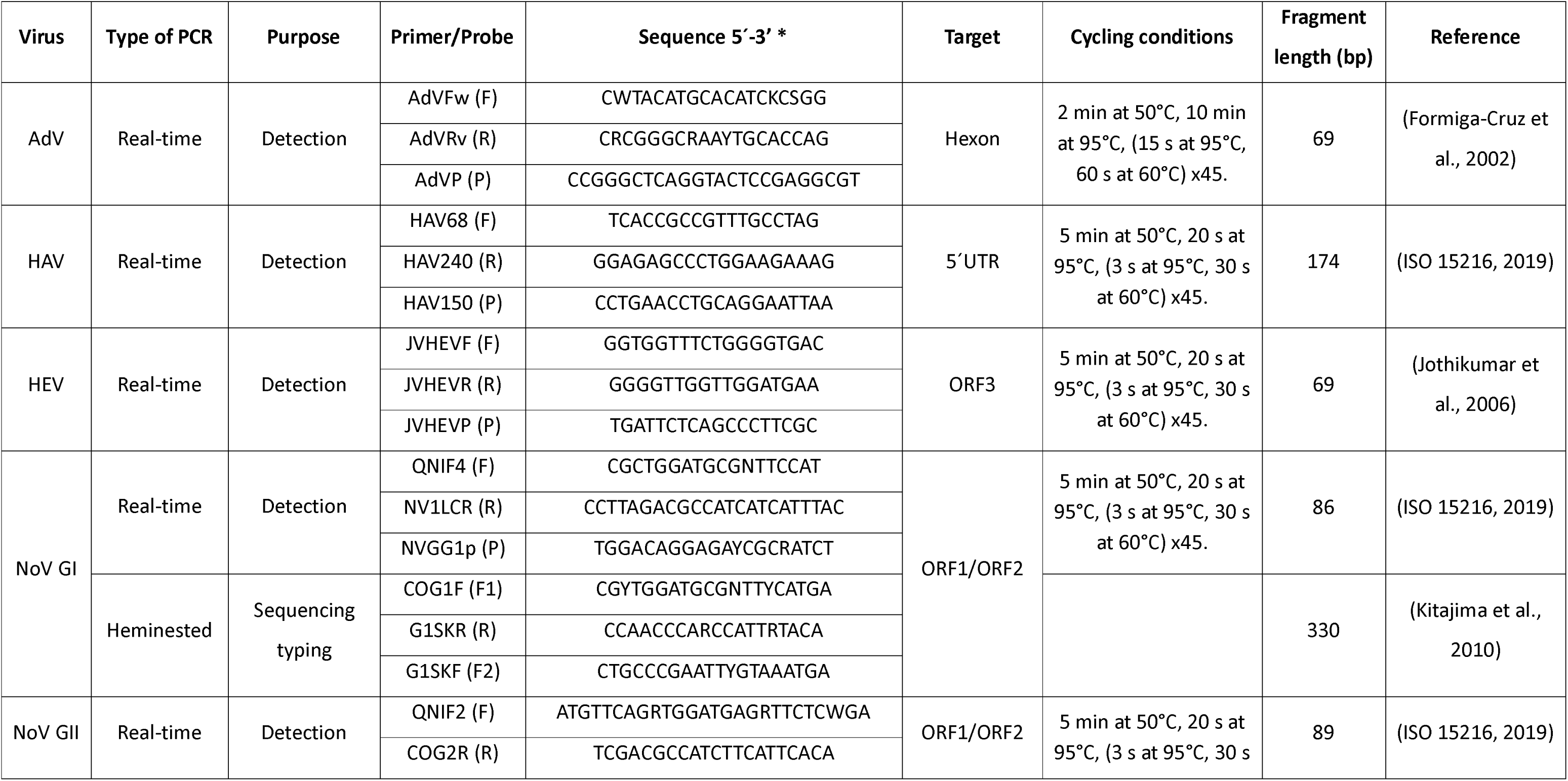
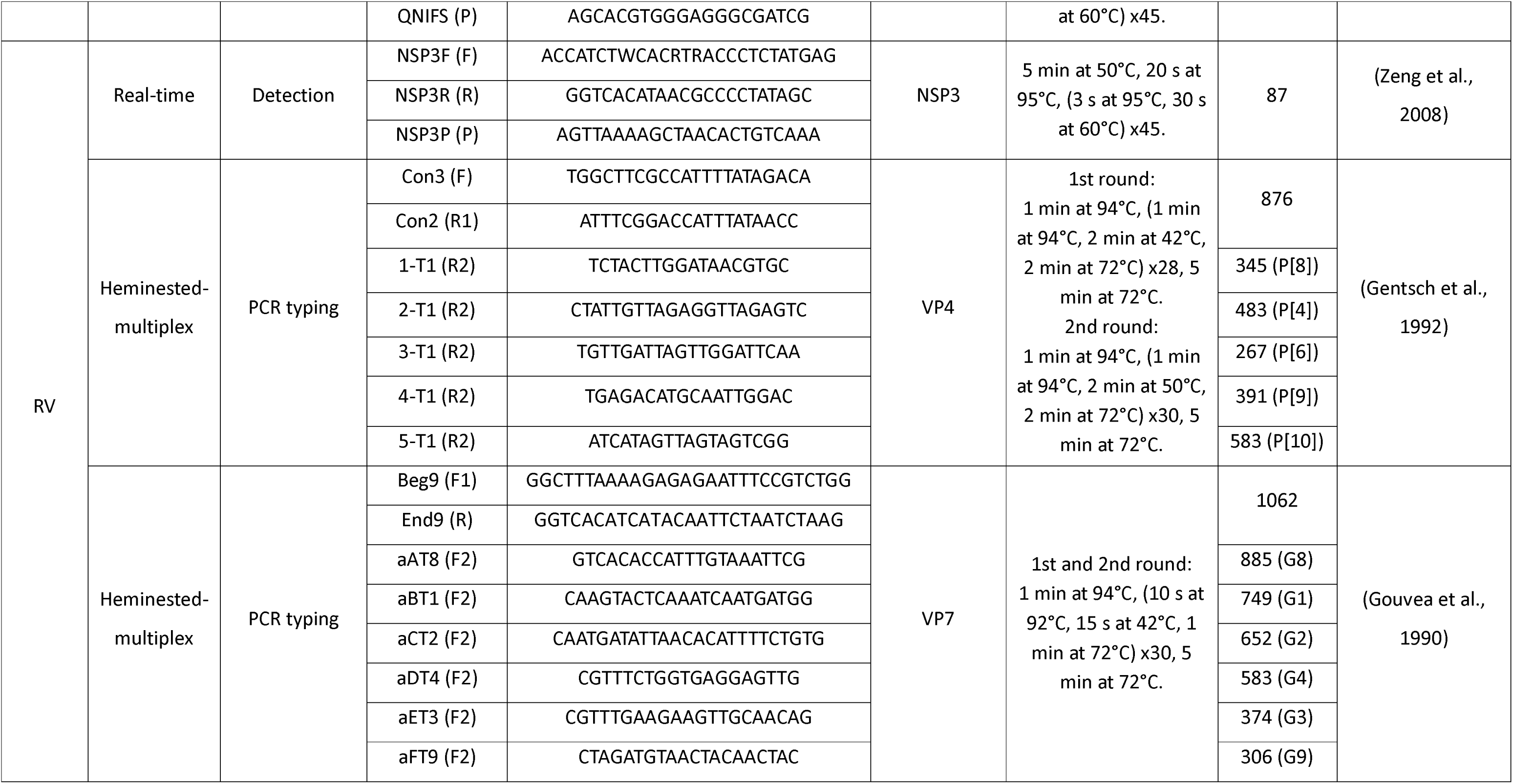
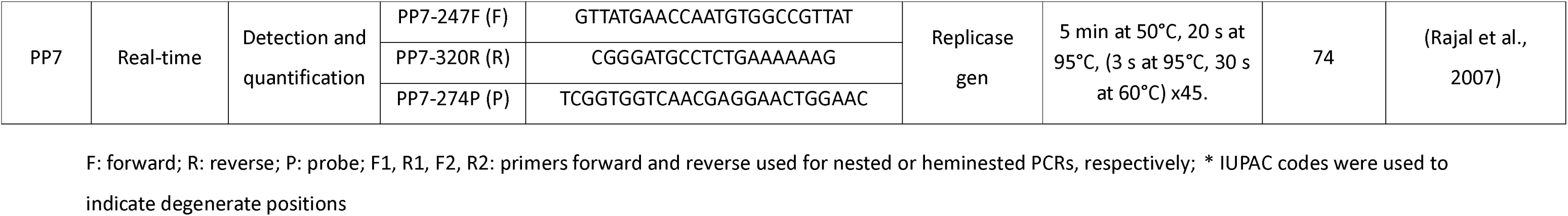
Primers and probes used for viral amplification.

For NoV, sequencing and phylogenetic analysis were performed to determine genotype. After amplification by RT-nested PCR using primers described in **Table 1**, the obtained PCR product was separated using 2% agarose gel electrophoresis and visualized with a UVP transilluminator. PCR fragment was isolated and purified using the PureLink® Quick Gel Extraction Kit (Invitrogen) and subjected to bidirectional Sanger sequencing using the ABI 3500xL Genetic Analyzer (Applied Biosystems). The resulting sequence was aligned and a phylogenetic tree was constructed from a dataset that included: 1) a sequence of each genotype, 2) the sequence obtained during this study, 3) the five most related strains obtained through BLAST (Basic Local Alignment Search Tool) analysis on the NCBI website (https://blast.ncbi.nlm.nih.gov/), 4) all previously obtained Argentine sequences (downloaded from GenBank), and 5) a NoV GII sequence as an outgroup. The maximum-likelihood (ML) method with IQTREE 1.6.12 (Trifinopoulos et al., 2016) employing the best-fit model of nucleotide substitution as determined by Model Finder (Kalyaanamoorthy et al., 2017) was used for phylogenetic analyses. The robustness of the clustered was assessed using the SH-like approximate likelihood ratio test (SH-like) with 1000 replicates (Guindon et al., 2010) and the ultrafast bootstrap (UFB) approximation with 10,000 replicates (Hoang et al., 2018). The obtained sequence was deposited in GenBank under the Accession Number PQ110318.

### 2.6 Statistical analyses

Fisher’s exact test was applied to evaluate differences in prevalence between fresh and frozen berries. Prevalences and their 95% confidence intervals were calculated using the *binom* package and the Pearson-Klopper method. All analyses were carried out in R version 4.0.5 (R Core Team, 2023).

## 3. Results

### 3.1 Virus detection in berries

Out of the total berries analyzed (n=145), 102 samples (70.3%) corresponded to frozen fruits and 43 (29.7%) to fresh fruits. Out of 145 samples, six (4.2%) tested positive for viral detection (three for NoV GI and three for RV), all of which were strawberry samples. Among the frozen fruits, 4.9% (5/102) tested positive, detecting NoV GI in three (2.9%) samples and RV in two (2.0%). For fresh fruit samples, one (2.3%) was positive for RV. No significant differences were found in prevalences between fresh and frozen fruits (p=0.67). No HAV or HEV were detected in any of the samples (**Figure 1 A**).

**Figure 1:**
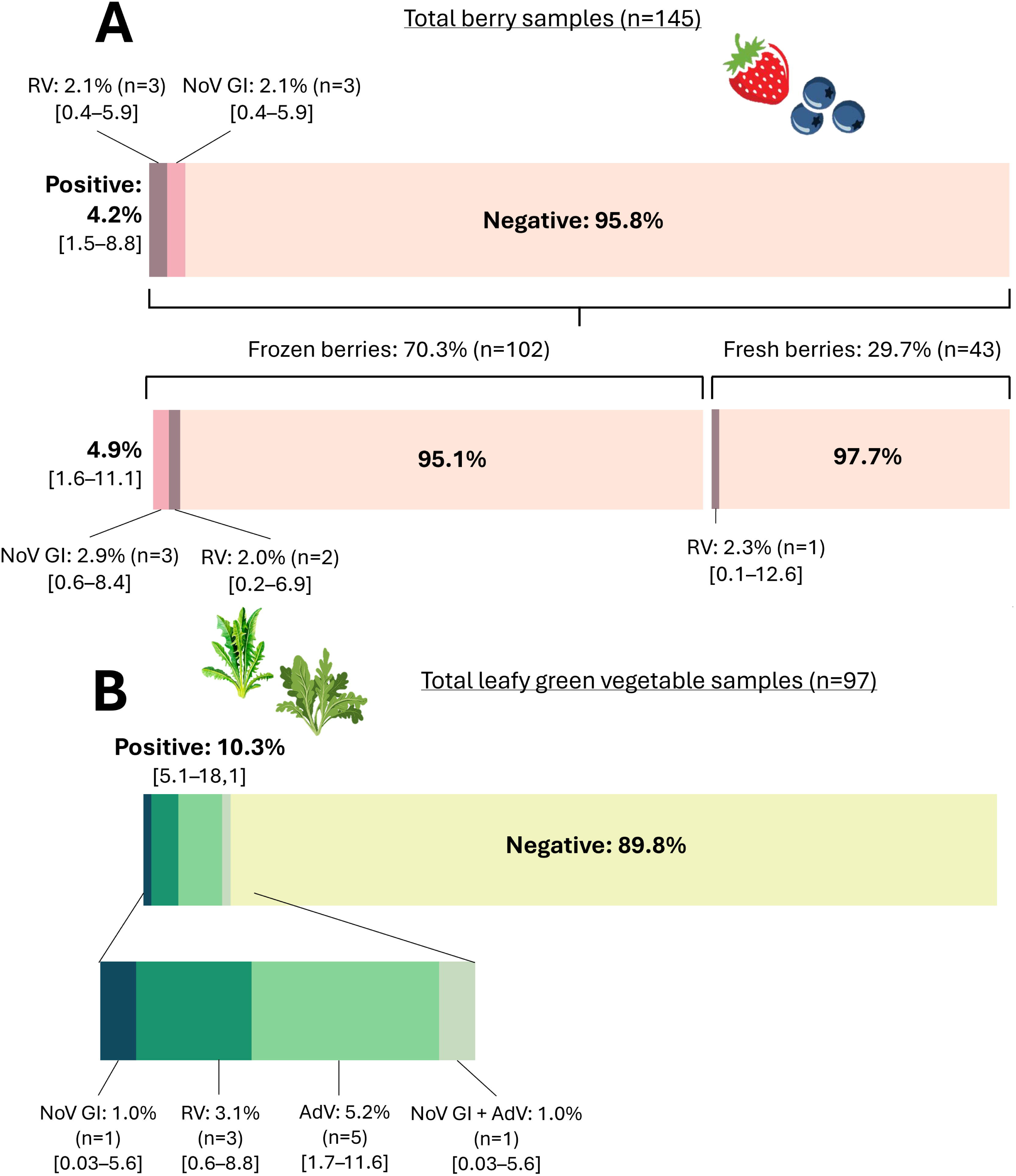
Viral molecular detection. The bars show the prevalence of viruses detected by real-time PCR in berries **(A)** and leafy green vegetables **(B)**.

PP7 was recovered from all viral concentrates (>1%). In instances where the recovery efficiency was less than 1%, 1/10 dilutions were conducted. Following dilution, PP7 was recovered with efficiencies exceeding 1% in all cases (as required by the ISO 15216-2:2019 standard).

One of the samples positive for NoV GI could be sequenced, and the results of the phylogenetic analysis showed that the sequence grouped with genotype 6 (**Figure 2**).

**Figure 2:**
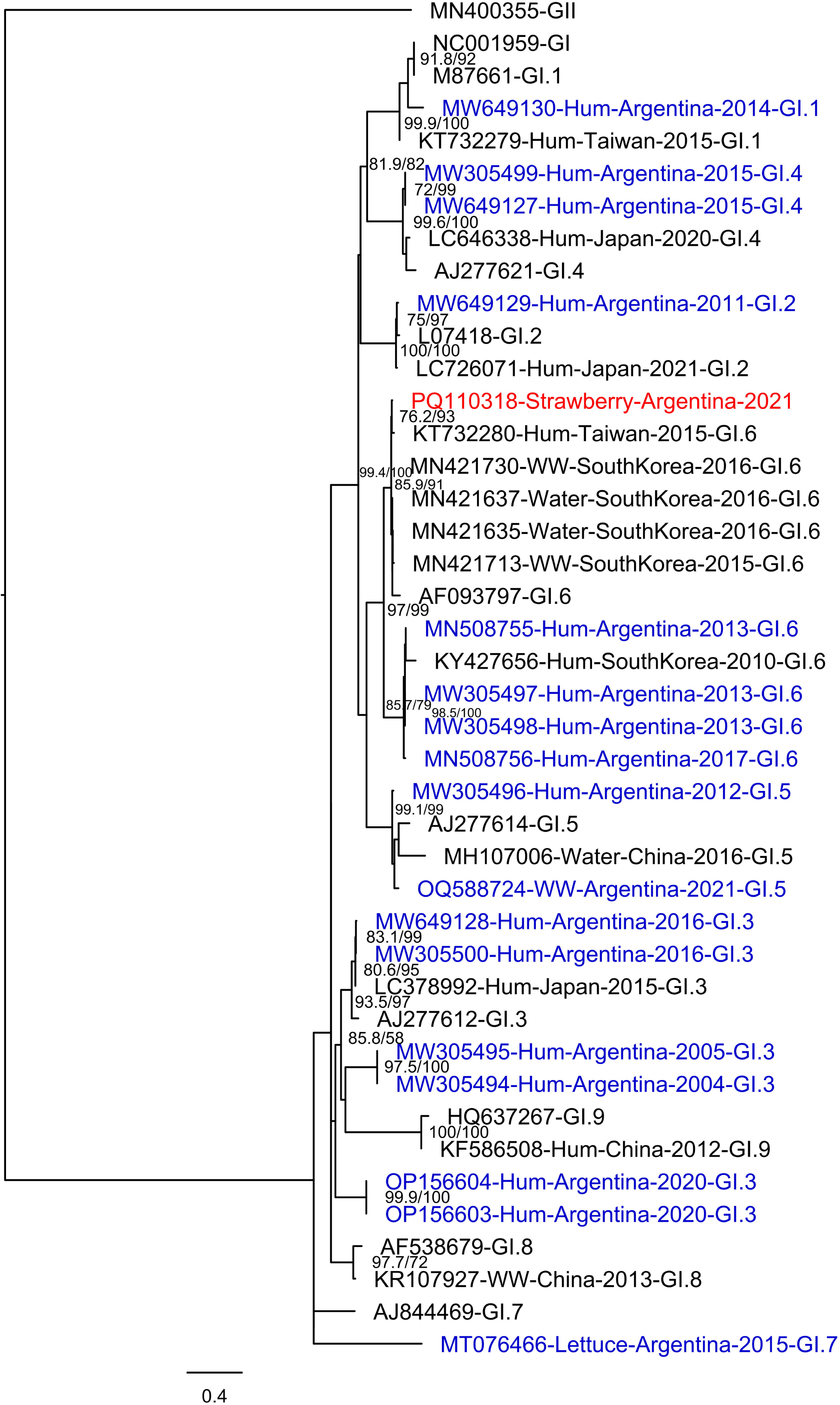
Phylogenetic tree obtained using the ML method with IQTREE 1.6.12 program, based on a 329 bp fragment of the ORF1/ORF2 of NoV. The sequence obtained in this study (red) were analyzed with Argentine sequences available in GenBank (blue) and two sequences of each genotype (black). Node supports: SH-like support (%)/UFB (%) (1000 and 10,000 replicates, respectively). Only supports over 70/70 are shown. References: WW= wastewater; Hum= clinical; Water= Tap water.

In one sample, genotype P[4] of RV was detected, while the other two samples could not be amplified for this region. Unfortunately, the G-types could not be identified.

### 3.2 Virus detection in leafy green vegetables

The overall virus detection rate was 10.3% (10/97), with nine positive cases found in rocket samples and one in chicory samples. Specifically, one sample tested positive for NoV GI (1.0%), three for RV (3.1%), five for AdV (5.2%) and one for both NoV GI and AdV (1.0%) simultaneously (**Figure 1 B**). The process control virus was successfully recovered from all viral concentrates with efficiencies surpassing 1%.

For RV, typing was successful in one of the samples, finding genotypes P[4] and P[10] along with genotype G2. However, the amplification of these regions could not be achieved in the two remaining samples. Unfortunately, the genomic fragments obtained for NoV GI and AdV could not be sequenced, likely due to low viral loads.

## 4. Discussion

Over the last year, data from the Ministry of Health of Argentina reported 44 outbreaks of foodborne diseases across the country, with the etiology confirmed in 24 of these outbreaks. Of the identified cases, 7 were of viral origin, primarily caused by NoV, followed by RV and AdV (Dirección De Epidemiología, 2025). However, it was not specified whether these outbreaks were linked to any particular food. This highlights the potential underestimation of foodborne viral transmission as a source of infection in our region.

In response to better understand viral contamination in frozen and fresh produce, we conducted a study to detect five viruses of human health significance in berries and RTE leafy vegetables collected in central Argentina. While some studies have explored enteric viruses in fresh produce, research specifically targeting RTE and frozen products in the Argentine market remains scarce. As emphasized by the FAO/WHO Expert Meeting, assessing enteric viruses in these foods and providing molecular data is essential for generating insights that can help prevent foodborne transmission. In our study, we successfully detected and characterized NoV, RV, and AdV, whereas HAV and HEV were not found.

### 4.1 Virus detection in berries

Our results showed the presence of NoV GI (2.1%) and RV (2.1%), primarily in frozen fruits. Genotyping revealed the presence of RV genotype P[4], while phylogenetic analysis of NoV revealed that the obtained sequence clustered with genotype 6 (GI.6), closely related to Argentine strains previously reported in association with gastroenteritis outbreaks (Degiuseppe et al., 2020; Tohma et al., 2021). These findings are novel for Argentina, as previous studies in berries reported the absence of noroviruses in strawberry samples collected in Buenos Aires province between 2014 and 2018 (Cammarata et al., 2021) or documented the presence of NoV GII with negative results for RV and HAV in berries collected from different regions of Argentina (Tucumán, La Pampa and Patagonia) in 2016, 2017 and 2020 (Oteiza et al., 2022).

Although no significant differences were observed between viral prevalences of fresh and frozen fruits, a higher tendency was noted in frozen ones, which could be attributed to better preservation of viruses at low temperatures (FAO/WHO, 2024). This finding is important, as frozen fruits are often consumed without prior washing, unlike fresh fruits, which are typically washed before consumption.

Several studies conducted worldwide have investigated enteric viruses in berries involved in outbreaks, showing high variability in both detection frequency and virus types. However, there are limited reports of viral screening in berry samples not directly associated with outbreaks; thus, there is scarce information regarding other viruses than NoV and HAV. Recent studies in berries from different countries have demonstrated a markedly reduced viral prevalence compared to previous studies conducted before 2011 (Miotti et al., 2024), with prevalence values lower than 3.6% (Cook et al., 2019; Oteiza et al., 2022; Steele et al., 2022; Terio et al., 2017; Torok et al., 2019), which coincides with our findings. This could be attributed to increased awareness of the impact of contaminated fresh or frozen produce on foodborne outbreaks, strict controls on berry exportation, advancements in agricultural practices, and improved hygiene and handwashing practices implemented in recent years. However, studies analyzing samples from Africa have reported higher viral prevalences (Miotti et al., 2024), likely due to poor hygiene and sanitary conditions in that region.

Given that berries produced in our country are primarily intended for export (RSA CONICET, 2016), either fresh or frozen, contamination by enteric viruses can potentially trigger foodborne outbreaks in distant regions of the world. Consequently, certain countries, such as the United States, Canada, and members of the European Union, request certification of berries entering their territories to be free of NoV and HAV (CBI, 2023), the primary causative agents of outbreaks. Due to international requirements, exported berries are tested for these two viruses, which contributes to their virological quality. However, in Argentina, there is no legislation requiring viral detection in berries intended for domestic consumption.

Furthermore, the detection of RV positive samples in this study highlights the importance of testing other pathogenic viruses in these foods.

### 4.2 Virus detection in leafy green vegetables

Regarding leafy green vegetables, we detected NoV, RV, and AdV in the absence of HAV and HEV. This is consistent with a meta-analysis, which described that AdV, RV, and NoV were detected more frequently than HAV and HEV (Barril et al., 2022). The finding of NoV GI is also in line with a recent meta-analysis, which reports that GI was more frequently detected in leafy vegetables worldwide compared to GII (J. Gao et al., 2024). Moreover, our detection of NoV GI and RV aligns with previous studies conducted in the central region of Argentina, which analyzed samples collected from retail sources or directly from cultivation sites and explored their correlation with irrigation water. These reports have shown frequent NoV and RV detection in vegetables cultivated in peri-urban areas of the city of Córdoba (central region of the country), along with analyses of the irrigation waters used in their cultivation (Coluccini, 2020; Prez et al., 2018, 2021). The investigations revealed that NoV was present in 58-60% of crop samples, with GI being the predominant genogroup, consistent with our findings. In those studies, RV was also detected in leafy vegetables, with prevalence ranging from 5% to 22% of samples, and the detected genotypes predominantly comprised G3 and G2, followed by G9, G4, and G1 for the VP7 gene, and P[8], P[10], P[4], P[6], and P[9] for the VP4 gene. In our study, we found genotypes P[4], P[10], and G2 in one positive sample. The detection of P[4] and G2 aligns with previous reports in the region and suggests that these genotypes have been circulating in the environment and food for at least eight years.

In this study, AdV was detected in 6.2% of the analyzed RTE samples. This is consistent with previous results reported in other regions of the world, such as Europe, South Korea or Africa (Cheong et al., 2009; Kokkinos et al., 2012; Traore et al., 2024), which documented prevalences between 6.7% and 26.4%. In South America, a recent study performed in southern Brazil also documented a comparable prevalence for AdV in lettuce samples (Gartner et al., 2022). To the best of our knowledge, this is the first study that reports AdV presence in fresh produce samples from Argentina, evidencing the presence of another gastroenteric virus that could potentially be transmitted to consumers.

The frequency of virus detection observed in this study was reasonably lower compared to the aforementioned reports conducted in central Argentina (Coluccini, 2020; Prez et al., 2018, 2021). This disparity could be attributed to the sanitization procedures employed for RTE leafy green vegetables, which involve washing with water and sodium hypochlorite. The effectiveness of disinfectants during RTE leafy greens production has been studied, and it has been concluded that washing with water or disinfectant solutions reduces natural microbial populations on the product’s surface (Allende et al., 2007; Beuchat et al., 2004). However, industrial disinfectant treatments do not guarantee the complete elimination of pathogens, particularly viruses (Terio et al., 2017), underscoring the potential for these RTE foods to serve as vehicles of pathogen transmission to consumers despite sanitation efforts.

### 4.3 Implications for human health

NoV has been identified as a major cause of gastroenteritis, responsible for about 200,000 deaths each year, predominantly in underdeveloped countries. NoV outbreaks are more frequent during winter due to its ability to thrive in cooler temperatures. Because NoV has high genetic variability, it has been challenging to develop effective interventions such as vaccines and antivirals (Winder et al., 2022). In Argentina, investigations of NoV foodborne outbreaks and routine diagnosis of acute gastroenteritis cases are infrequent. In our study, NoV was detected in both types of food tested, suggesting that its presence in fruits and vegetables may be common in central Argentina.

RV is a significant pathogen responsible for diarrheal illnesses in both humans and animals. It is widely recognized as a leading cause of acute gastroenteritis in young children worldwide (Sadiq et al., 2018). Many countries have observed a reduction in rotavirus disease burden following the introduction of rotavirus vaccines (Crawford et al., 2017). In Argentina, the monovalent Rotarix™ vaccine (G1P[8]) has been included in the national vaccination program since 2015, aiming to reduce morbidity and mortality, although it does not prevent infection. This vaccination strategy has had a positive impact, leading to a decline not only in specific rotavirus cases but also in overall acute diarrhea cases in Argentina (Degiuseppe & Stupka, 2018). Vaccination has also been associated with changes in circulating rotavirus genotype patterns, with an increased detection of genotypes G2P[4], followed by G3P[8] and G9P[8] in clinical cases (Degiuseppe & Stupka, 2018). In this study, RV was detected in both strawberries and rocket, indicating its circulation within the country and the potential risk of transmission through these foods. The P[4] genotype, reported as the most widely circulating in humans in Argentina during 2021 and 2022 (Degiuseppe et al., 2023), was detected in both sample types, suggesting its broad distribution across various food matrices. Conversely, there have been no recent reports of P[10] detection in clinical, environmental or food samples in Argentina, which provides insight into the circulation of an unusual genotype in the region.

AdV constitutes a group of viruses responsible for a wide range of infections, which are commonly excreted by human populations worldwide and are often found in high concentrations in wastewater (Formiga-Cruz et al., 2002; Pina et al., 1998). Consequently, they are considered suitable indicators of fecal contamination in both drinking water and food (Kokkinos et al., 2012). During this investigation, AdV was found in leafy green vegetable samples, which could indicate fecal contamination (e.g. through contaminated irrigation waters) and/or poor food handling, as contamination may occur at different stages of the production chain (Hewitt et al., 2013).

HAV and HEV are acute hepatitis-producing agents that circulate worldwide. HAV infects only humans, while HEV is a zoonotic virus with pigs, wild boars, and deer as its main reservoirs (Pisano et al., 2018; Ré et al., 2024). Although both viruses have been reported in Argentina and previously identified in humans, pigs (only HEV), and environmental samples such as wastewater and dam water (Di Cola et al., 2023; Fantilli et al., 2023, 2024; Martínez Wassaf et al., 2014; Masachessi et al., 2018), neither virus was detected the analyzed food samples. This finding suggests that foodborne transmission may not constitute a significant source of infection for these viruses in Argentina. However, a larger number of samples must be studied to confirm this hypothesis. Therefore, despite their documented presence in the region, the absence of HAV and HEV in berries and RTE vegetables indicates a lower level of circulation, particularly for HAV. This reduced prevalence could be attributed to the national HAV vaccination program implemented in Argentina since 2005, in contrast to the higher detection rates observed for NoV, RV, and AdV.

While the detection of viruses through molecular methods does not necessarily demonstrate that these foods can cause infections in consumers, it suggests inadequate decontamination, processing measures, handling, or storage practices of RTE products. Detecting the viral genome does not implicate viable viral particles, but it clearly indicates their previous presence (Bosch et al., 2018). Due to the challenges associated with the use of cell culture for assessing the viability of enteric viruses (Ha et al., 2016; Papafragkou & Kulka, 2016), several experimental approaches have been developed to date to differentiate infectious from non- infectious viruses using modified molecular amplification (e.g., coupling with propidium monoazide to selectively block DNA/RNA from non-viable viruses). However, a standardized method to definitively assess viral infectivity has yet to be established (FAO/WHO, 2023).

Considering that enteric viruses are highly resilient to environmental conditions and that RTE foods do not undergo any subsequent treatment to ensure virus inactivation, there is an increased likelihood of infection transmission to consumers, highlighting the need for stringent food safety measures throughout the production and distribution chain. The findings from this study revealed viral contamination in RTE fresh and frozen produce, underscoring the importance of strengthening post-harvest hygiene protocols to reduce viral transmission and prevent foodborne outbreaks. Our results contribute to the knowledge needed to improve access to safe and healthy food and enhance consumer protection, in line with the Sustainable Development Goals (SDG) 3 (Good Health and Well-being) and SDG 12 (Responsible Consumption and Production) of the WHO 2030 Agenda for Sustainable Development. These findings present new challenges for food control systems, urging public policymakers to promote a food safety culture by establishing evidence-based regulations that include virus monitoring in foods, especially given that certification of the absence of NoV and HAV is currently required for food exports to certain countries but not for those intended for domestic consumption.

## Data Availability

All data produced in the present work are contained in the manuscript

## Author contributions

**Guadalupe Di Cola:** Conceptualization, Methodology, Investigation, Writing - Original Draft**; Verónica E. Prez:** Conceptualization, Methodology, Investigation**; Anabella C. Fantilli:** Methodology, Investigation**; Camila Frydman:** Methodology**; Marina V. Mozgovoj:** Methodology, Visualization**; Liliana Luque:** Methodology**; Leonardo Ferreyra:** Methodology, Visualization**; Silvia V. Nates:** Conceptualization, Visualization, Funding acquisition**; María Belén Pisano:** Conceptualization, Writing - Review & Editing, Supervision, Funding acquisition**; Viviana E. Ré:** Conceptualization, Writing - Review & Editing, Supervision, Funding acquisition.

